# The role of school enjoyment in the association between externalising and depressive symptoms and academic attainment: findings from a UK prospective cohort study

**DOI:** 10.1101/2020.05.21.20108928

**Authors:** Tim Cadman, Amanda Hughes, Caroline Wright, José A López-López, Tim Morris, Frances Rice, George Davey Smith, Laura D Howe

## Abstract

Previous research on the relationship between children’s externalising and depressive symptoms, experience of school, and later academic attainment is inconclusive. The present study uses data from the Avon Longitudinal Study of Parents and Children (n=6,409) to investigate bidirectional associations between school experience (enjoyment and connectedness) and externalising and depressive symptoms at age 10-11 and 13-14. We also investigate the relationship between school experience and academic attainment at 16 and test whether school experience mediates associations of externalising and depressive symptoms with later attainment. A cross-lagged structural equation model was employed. Externalising and depressive symptoms at 10-11 were negatively associated with school connectedness at 13-14 (externalizing: standardised β=−0.13, CI: −0.17, −0.08; depressive: β=−0.06, CI: −0.11, 0.01), and with school enjoyment at 13-14 (externalising: β=−0.08, CI: −0.13, −0.03; depressive β=−0.04, −0.08, 0.03). School enjoyment at 13-14 was positively associated with attainment at 16 (β=0.10, CI: 0.04, 0.15), and partially mediated associations between externalising and depressive symptoms at 10-11 and attainment at 16 (externalising: proportion mediated; 4.7%, CI: 0.7, 10.1, depressive: proportion mediated 2.2%, I: −1.5, 5.9). School enjoyment is a potentially modifiable risk factor that may affect educational attainment of adolescents with depressive and externalising symptoms.

---

There is consistent evidence that children with hyperactive and conduct difficulties (1-8) and depression (4, 5, 7) have on average lower academic performance than unaffected peers. This is of importance because poorer educational attainment in childhood is linked with worse health and economic outcomes in adulthood. (9) To improve life-long outcomes for children with these difficulties it is necessary to understand the relationship between these problems and attainment and identify modifiable mediating factors.

One potential mediator is children’s experience and enjoyment of school. School climate in the UK – which emphasises quiet, focused work – can be a poor ‘fit’ for children with attentional and behavioural problems. (10, 11) Children with these difficulties elicit more negative responses from staff, have poorer relationships with peers and are more likely to be excluded from school. (12-14) Adolescent depression is also associated with poorer social functioning, (15) friendships (16) and academic self-concept, (17) which could result in lower school enjoyment. There is also growing evidence that school enjoyment may positively relate to academic outcomes. (18-21)

Longitudinal research on the relationship between school experience and externalising and depressive symptoms is limited. Whilst positive prospective associations between school experience and externalising and depressive symptoms have been consistently reported, (22-29) evidence for the converse associations - between externalising / depressive symptoms and school experience - is inconsistent. Using a cross-lagged design Lester et al. found bidirectional associations between depression and anxiety and children’s feeling of connection with school, (30) but Shochet et al. reported the association between anxiety and depressive symptoms and school experience to be close to null. (27) A third study by Loukas et al (n=296) was too small to draw any firm conclusions. (31) Importantly, there is emerging evidence that both children’s school experience and their socioemotional well-being can be improved by school-based interventions. For example, a recent RCT in India reported that a school-based intervention which focused on teaching problem-solving skills and engaging students and teachers in decision-making improved student’s perceptions of their school and reduced rates of depressive symptoms. (32) Positive results from similar interventions have been reported in the UK and USA. (33, 34)

The current study had three main aims: (i) to test the associations between externalising / depressive symptoms and school enjoyment at two time points (ages 10-11 and ages 13-14), (ii) to explore associations between school experience at ages 13-14 and academic attainment age 16, and (iii) to test whether school experience at ages 13-14 mediates the relationship between externalising or depressive symptoms at age 10-11 and academic attainment at age 16.

## METHODS

### Sample

Data was used from the Avon Longitudinal Study of Parents and Children (ALSPAC). Pregnant women resident in Avon, UK with expected dates of delivery 1st April 1991 to 31st December 1992 were invited to take part in the study. The initial number of pregnancies enrolled is 14,541 (for these at least one questionnaire has been returned or a “Children in Focus” clinic had been attended by 19/07/99). Of these initial pregnancies, there was a total of 14,676 foetuses, resulting in 14,062 live births and 13,988 children who were alive at 1 year of age. (For further details on the cohort profile, representativeness, and phases of recruitment, see (35, 36). The study website contains details of all data available through a fully searchable data dictionary and variable search tool: http://www.bristol.ac.uk/alspac/researchers/our-data/. Ethical approval for the study was obtained from the ALSPAC Ethics and Law Committee and the Local Research Ethics Committees. Informed consent for the use of data collected via questionnaires and clinics was obtained from participants following the recommendations of the ALSPAC Ethics and Law Committee at the time.

### Measures

#### Externalising

Externalising symptoms were measured via maternal report using the Conduct and Hyperactivity subscales of the Strengths and Difficulties Questionnaire at mean ages 11.5 (T1) and 13 (T2). (37) Both subscales contain 5 items scored 0, 1 or 2 giving a total range of 0 – 10, with higher scores indicating greater difficulties. These scales were modelled as one factor in the main analysis, but additional analyses modelled the scales separately.

#### Depressive symptoms

Depressive symptoms were self-reported using the Short Mood and Feelings Questionnaire at mean ages 10.5 (T1) and 14 (T2). (38) This questionnaire contains 13 items scored 0, 1 or 2 giving a total range of 0 – 26, with higher scores indicating greater depressive symptoms.

#### School experience

School experience was assessed via self-report at ages 11 (T1) and 14 (T2). Previous research has posited multiple overlapping constructs related to school experience, (39) here we focus on two aspects: (i) enjoyment and (ii) connectedness. (26) School enjoyment items refer to the child’s enjoyment of classes and school (for example “School is a place where I enjoy what I do in class”). School connectedness items ask about the child’s sense of belonging at the school and how positively they believe they are viewed by others (for example “School is a place where people can depend on me”), and is closely related to concepts such as school climate. (27, 32) Higher scores indicate greater enjoyment and connection. Full details of questions are provided in Tables S2 & S4.

#### Academic attainment

Academic attainment was measured by capped General Certificate of Secondary Education (GCSE) point score at age 16 (T3), obtained via linkage to the National Pupil Database. GCSEs are qualifications taken at age 16, which represented the end of compulsory education in the UK for this cohort. Capped GCSE point scores are continuous scores representing the best 8 grades at GCSE. Each grade has a different value (for example A* which represents the highest possible grade is worth 58 points, C is worth 40 points) and these are summed to produce the total score. (40) For the majority of students the capped score will range between 0 – 464; however it is possible some may have scores up to 540 if they took more advanced qualifications than GCSE at age 16. The capped, rather than total GCSE score was chosen because it reduces the influence of scores from pupils who take more than the standard number of examinations.

#### Covariates

The following variables were identified *a priori* as confounders as they had either previously been found to or could plausibly influence externalising, depressive symptoms, school experience and attainment: maternal smoking in pregnancy (yes/no), housing tenure (owner/mortgaged vs other), highest parental education (None, Vocational, O-level, A-level, Degree), self-reported material hardship (yes/no), parity, IQ and earlier school enjoyment. Early school enjoyment was measured by binary items at ages 5 and 6 asking the child whether they enjoy going to school. Child IQ was assessed at age 8 using a short form of the Weschler Intelligence Scale for Children - III, (41) whilst all other confounders were measured in pregnancy. We adjusted for child sex at birth as it is associated with externalising, depressive symptoms and attainment and this adjustment may have improved the precision of results.

### Statistical analysis

The relationship between study variables was tested using structural equation modelling. Depressive symptoms, externalising, school enjoyment and school connectedness were modelled as latent variables. A cross-lagged panel design was used to test the associations between school experience and externalising/depressive symptoms at ages 10-11 and 13-14 (Figure 1; separate models for externalising and depressive symptoms). The cross-lagged design allows the estimation of associations between multiple time points whilst controlling for within time-point correlations and the stability of constructs over time. (42) Additional correlations were specified between the error terms of pairs of items repeated at T1 and T2. All results are presented in standardised coefficients (β) to allow comparison between estimates.

**Figure 1:**
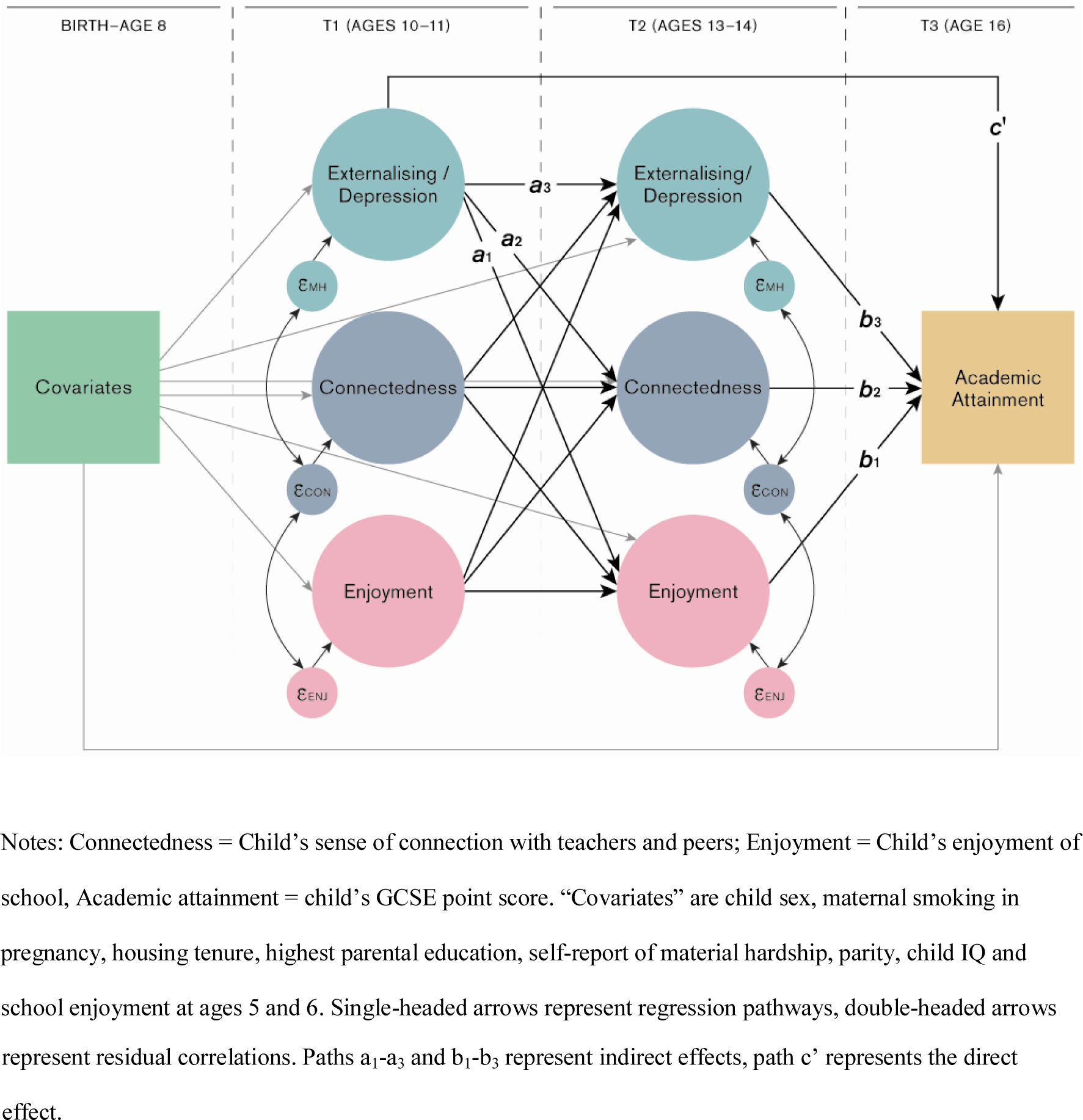
Structural equation model diagram.

Potential mediation of the association between externalising/depressive symptoms and academic attainment via school experience was also tested within the structural equation modelling framework. Three indirect effects (Figure 1: a_1_ × b_1_, a_2_ × b_2_ and a_3_ × b_3_) were calculated from the paths from age 10-11 externalising/depressive symptoms to age 16 attainment. The direct effect (c’) is the association between age 10-11 externalising/depressive symptoms and age 16 attainment whilst adjusting for age 13-14 externalising/depressive symptoms, enjoyment and connectedness.

For both depressive and externalising symptoms, an unadjusted model was tested followed by two levels of adjustment (i) a minimally adjusted model (child sex, maternal smoking in pregnancy, housing tenure, highest parental education, self-report of material hardship, parity and school enjoyment at ages 5 and 6) and (ii) a model additionally adjusted for child IQ. As previously argued, (6) although IQ is potentially a confounder of the relationships between externalising/depressive symptoms, school experience and attainment, it may also be on the causal pathway from earlier externalising/depressive symptoms or school experience to later attainment. If differences in IQ are (even partially) caused by earlier externalising/depressive symptoms or school experience (e.g. through lower motivation or concentration), then adjusting for IQ would bias the estimates downwards. Conversely, if IQ is purely a confounder then an IQ-adjusted model would be correct. However, given the available data it is not possible to distinguish between the two situations. The main results are presented for the minimally adjusted model, with all models reported in Supplementary Materials. Child sex was associated with most exposures and outcomes (Table S3) therefore analyses were also repeated stratified by sex. The model was estimated using Diagonal weighted least squares with robust versions of fit statistics, using the lavaan package in R version 3.53.

### Inclusion criteria, and method for addressing missing data

Missing data were imputed for all variables in the model using multiple chained equations for the sample of subjects with complete data on GCSE attainment at age 16 and at least one measure of externalising or depressive symptoms and one school experience measure at one time point. Data were imputed separately for males and females and then combined. The imputation model contained all variables within the model and additional variables known to be predictors of missingness including earlier measurements of school enjoyment and externalising/depressive symptoms (full details of the imputation model and distribution of original and imputed data are provided in Supplementary Materials). 20 datasets were imputed and estimates combined using Rubin’s rules. (43)

## RESULTS

### Descriptive statistics

The mean GCSE point score was 344.67 (SD: 78.83). Mean IQ score at age 8 was 103.88 (SD: 16.42), indicating scores slightly above the standardised average of 100. Distributions of school experience variables were skewed towards a positive school experience, and distributions of depressive symptoms and externalising towards lesser symptoms (Table S2).

Correlations between school enjoyment and connection were 0.73 (95% confidence interval (CI): 0.72, 0.74) at ages 10-11 and 0.58 (CI: 0.56, 0.60) at ages 13-14. Correlations between externalising and depressive symptoms were 0.24 (CI 0.20, 0.27) at ages 10-11 and 0.21 (CI: 0.17, 0.25) at ages 13-14.

Descriptive statistics suggest that the analysis sample was of higher average socioeconomic position compared to the full ALSPAC sample (Table S2). For example, mean GCSE point score in the full ALSPAC sample was 318.94 (SD: 93.95) compared to 344.67 (SD: 78.33) in the analysis sample; similarly average maternal age at birth in full ALSPAC sample was 28.64 (SD: 4.86) vs 29.31 (SD: 4.55) in the analysis sample.

### Model fit

Fit for the latent models of school experience, externalising and depressive symptoms was good (average fit statistics across imputations, externalising model RMSEA = 0.06, SRMR = 0.06, CFI = 0.97, TLI = 0.97, depressive symptoms model RMSEA = 0.04, SRMR = 0.04, CFI = 0.99, TLI = 0.99). Loadings were moderate to high on all items (0.43 – 0.95), apart from one item (“School is a place where I get excited about the work I do”) with a very low loading of 0.01 (Tables S2 - S4). This item was retained in the model to allow comparison with previous findings from the ALSPAC cohort.

### Main findings

The key results are shown in Figures 2 and 3 and Tables S9 and S10.

**Figure 2:**
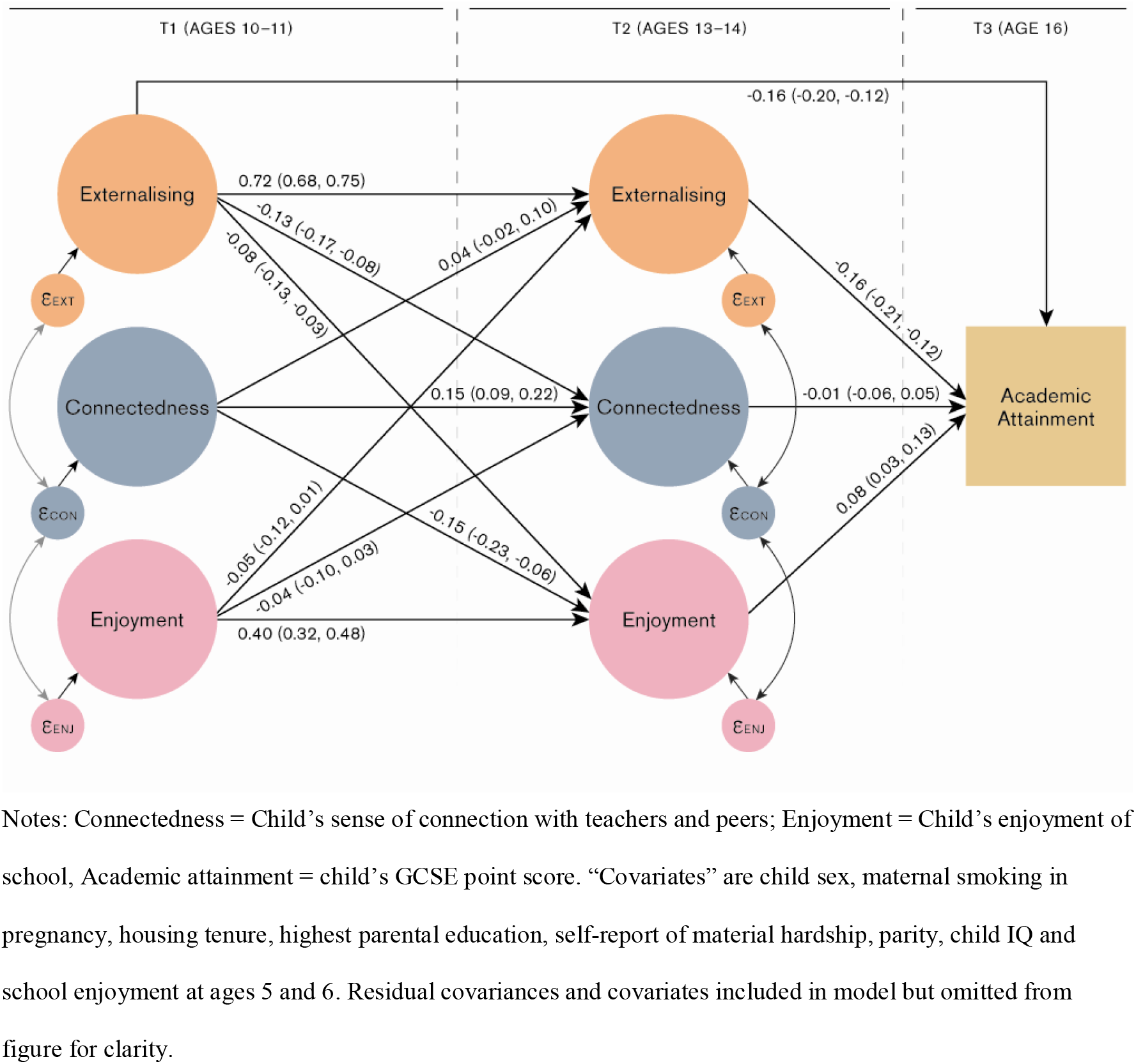
Cross-lagged model of depressive symptoms and school experience (n=6,409)

**Figure 3:**
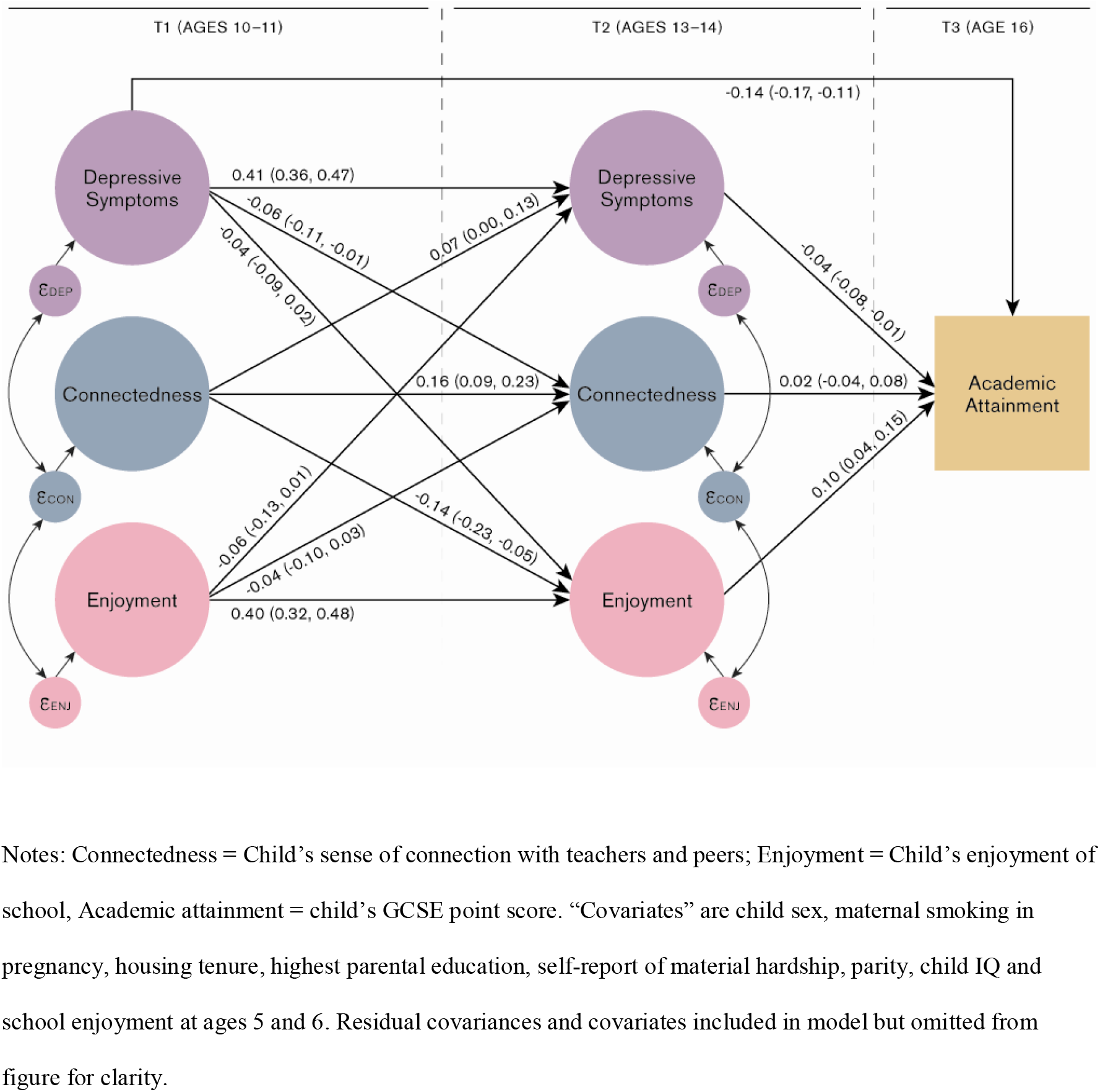
Cross-lagged model of depressive symptoms and school experience (n=6,409)

#### Stability of school enjoyment, externalising and depressive symptoms over time

School enjoyment showed moderate stability between ages 10-11 and 13-14 (β = 0.40, CI: 0.32, 0.48) whilst connectedness showed lower stability (β = 0.15, CI: 0.09, 0.22). Externalising showed the highest stability between time points (β = 0.72, CI: 0.68, 0.75) whilst the stability of depressive symptoms was lower (β = 0.41, CI: 0.36, 0.47).

#### Associations between school experience and externalising symptoms at ages 10-11 and 13-14

There was weak evidence of a negative association between school enjoyment aged 10-11 and externalising at age 13-14 (β = −0.05, CI: −0.12, 0.01). We found also found weak evidence for a positive association between school connectedness age 10-11 and externalising age 13-14, though confidence intervals crossed the null β = 0.04, CI: −0.02, 0.10).

There was clearer evidence of negative associations between externalising age 10-11 and school experience at 13-14 (externalising to enjoyment: β = −0.08, CI: −0.13, −0.03; externalising to connectedness: β = −0.13, CI: −0.17, −0.08). Modelling hyperactivity and conduct problems separately indicated stronger associations with school experience age 13-14 for conduct problems than for hyperactivity/impulsivity at 10-11 (Tables S17 & S18). Associations were similar for males and females, except for that of age 10-11 externalising symptoms with age 13-14 enjoyment which was larger for boys than girls. (Tables S12 & S13). Associations did not differ notably when additionally adjusted for IQ.

#### Associations between school experience and depressive symptoms at ages 10-11 and 13-14

A similar pattern of results was found for depressive symptoms. There was weak evidence of a negative association between school enjoyment age 10-11 and depressive symptoms age 13-14 (β = −0.06, CI: −0.13, 0.01). We also found evidence for a positive association between connectedness age 10-11 and depressive symptoms age 13-14 (β = 0.07, CI: 0.00, 0.13).

Negative associations were observed between depressive symptoms age 10-11 and school experience age 13-14 (depressive symptoms to enjoyment: β = −0.04, CI: −0.09, 0.02; depressive symptoms to connectedness: β = −0.06, CI: −0.11, −0.01). Analyses stratified by sex indicated that associations were larger for girls than boys (Tables S13 & S14). There was only minimal attenuation of associations after additionally adjusting for IQ.

#### Associations between externalising/depressive symptoms age 13-14 and academic attainment age 16

Externalising symptoms at both ages 10-11 and 13-14 were negatively associated with attainment at age 16 (age 10-11: β = −0.16, CI: −0.21, −0.10; age 13-14: β = −0.16, CI: −0.20, −0.12), as were depressive symptoms (age 10-11: β = −0.14, CI: −0.17, −0.11; age 13-14: β = −0.04, CI: −0.08, −0.01). These associations were attenuated by approximately 25-50% once adjusting for IQ.

#### Associations between school experience age 13-14 and academic attainment age 16

After adjusting for potential confounders, school enjoyment at age 13-14 was positively associated with attainment at age 16 (β = 0.08, CI: 0.03, 0.13); but connectedness was not (β = −0.01, CI: −0.06, 0.05). Adjusting for child IQ did not substantially change these estimates. The association between school enjoyment and attainment was notably larger for girls than boys (girls β = 0.15, CI: 0.09, 0.21; boys β = 0.03, 95% CI: −0.14, 0.20)

#### Does school experience mediate the association between externalising /depressive symptoms and attainment?

There was evidence that school enjoyment partially mediated the association between externalising and depressive symptoms ages 10-11 and attainment at age 16 (Figure 1, path a_1_ × b_1_). The proportion of the association between age 10-11 externalising and age 16 attainment mediated via age 13-14 enjoyment was 4.7% (CI: 0.7, 10.1); for depressive symptoms to attainment the proportion mediated via enjoyment was 2.2%, (CI: −1.5, 5.9). However, there was minimal evidence of mediation via school connectedness (Figure 1, path a_2_ × b_2_, proportion mediated externalising to attainment via connectedness −0.5%, CI: −5.2, 4.3; proportion mediated depressive symptoms to attainment via connectedness 0.6%, CI: −1.6, 2.8).

## DISCUSSION

The aims of this study were: (i) to test associations between externalising/depressive symptoms and school experience at two time points in secondary school (ages 10-11 and ages 13-14), (ii) to explore whether school experience at ages 13-14 was associated with academic attainment age 16, and (iii) to test whether school experience at ages 13-14 mediated the relationship between externalising/depressive symptoms at age 10-11 and academic attainment at age 16.

Results support negative associations between externalising and depressive symptoms at age 10-11 and school connectedness at age 13-14. There was weaker evidence for associations between externalising and depressive symptoms age 10-11 and school enjoyment age 13-14. We found some evidence for associations between school experience age 10-11 and externalising and depressive symptoms age 13-14, though confidence intervals crossed the null. School enjoyment at age 13-14 was positively associated with attainment at age 16, and partially mediated the relationship between earlier externalising and depressive symptoms and attainment. Although the magnitude of associations was modest, our findings suggest that children with symptoms of conduct problems, inattention/hyperactivity and depression may find school less enjoyable after the transition to secondary school, which in turn affects their later academic attainment.

### Externalising and school experience

The negative associations between externalising symptoms at age 10-11 and connectedness aged 13-14 indicates that an aspect of school experience most affected for children with externalising difficulties is their relationships with teachers and pupils. This complements existing findings that externalising negatively impacts on peer relations, (12, 13) which may in turn may be one pathway to elevated levels of depression. (44, 45) We found some evidence that conduct problems rather than hyperactivity were more strongly associated with subsequent school enjoyment; however further studies are needed to replicate this finding.

Reduced school enjoyment and connectedness for children with attentional and behavioural problems likely results from a complex interplay of individual and societal factors. School culture in the UK rewards quiet, focused and obedient behaviour, (10) and can be a particularly poor fit for children with attentional and behavioural difficulties. (11) Children entering secondary school with these difficulties are likely to struggle to meet many of the demands of school which in turn could create feedback loops.(46) Their behaviours may elicit a negative reaction from peers and teachers (12, 13) which in turn could result in an escalation of behaviours, leading to an increased risk of exclusion and absence, (14, 47) low mood and poor academic outcomes. (19-21)

### Depressive symptoms and school experience

Similarly to externalising, children suffering depressive symptoms at the transition to secondary school were less likely to feel connected to teachers and peers at age 13-14. This fits with previous findings that depression is associated with impaired relationships. (15, 17) The association between depressive symptoms and school enjoyment may also reflect a cognitive bias towards negative evaluation amongst children with high levels of depressive symptoms. (48) One puzzling finding was a *positive* association between school connectedness age 10-11 and depression and externalising age 13-14. However, this association only emerged when conditioning on school enjoyment.

### School experience, externalising/depressive symptoms and attainment

Consistent with previous research, (18-21) we found a positive association between school enjoyment at age 13-14 and attainment at age 16, after controlling for externalising/depressive symptoms and potential confounders. However we found close to no association between connectedness age 13-14 and attainment age 16. One explanation of this difference is that two of the three items relating to enjoyment specifically ask about enjoying work, which may be more relevant to future attainment than relationships with teachers and peers. We also found notable sex differences, with a stronger association between school enjoyment and academic outcomes for boys than girls. Our finding that school enjoyment partially mediates the relationship between externalising/depressive symptoms and academic attainment suggests children’s experience of school may be a potentially modifiable risk factor, although the proportion mediated was small.

Importantly, there is evidence that school enjoyment and connectedness is modifiable. Whilst there is limited evidence for the effectiveness of individual interventions to improve children’s emotional well-being, (11, 49) trials using whole-school approaches have shown promising results. (32, 33, 50) For example, a recent RCT in India found that an intervention which taught problem-solving skills and engaged students and teachers in decision-making reduced rates of depressive symptoms and improved students’ perception of the school climate. (32) Positive results have also been reported in UK and US, (33) (50) though further high-quality trials are needed.

### Limitations

This was a large prospective cohort study with repeated data on externalising, depressive symptoms and school experience along with linked educational data. The study design attempted to ascertain the direction of relationships and we were able control for known confounders including child IQ. Nevertheless, as an observational study, results will be subject to residual confounding and may suffer from reverse causation. Attrition in ALSPAC is high and associated with sex and socioeconomic factors, (51) and whilst multiple imputation was used to mitigate resultant bias our imputed sample remained of higher socioeconomic position than the full ALSPAC sample.

### Summary

In summary, we found evidence that externalising and depressive symptoms were prospectively associated with school connectedness, with weaker evidence for other pathways. School enjoyment was associated with academic attainment, and may partially mediate associations between externalising and depressive symptoms and later academic attainment, indicating a possible avenue for intervention.

## Data Availability

Data available from authors on request.

## ACKNOWLEDGEMENTS

We are extremely grateful to all the families who took part in this study, the midwives for their help in recruiting them, and the whole ALSPAC team, which includes interviewers, computer and laboratory technicians, clerical workers, research scientists, volunteers, managers, receptionists and nurses.

The UK Medical Research Council and Wellcome (Grant ref: 217065/Z/19/Z) and the University of Bristol provide core support for ALSPAC. This publication is the work of the authors and will serve as guarantors for the contents of this paper. A comprehensive list of grants funding is available on the ALSPAC website (http://www.bristol.ac.uk/alspac/external/documents/grant-acknowledgements.pdf); This research was specifically funded by the UK Medical Research Council (MR/M020894/1) and the Health Foundation’s Social and Economic Value of Health Programme (Grant ID: 807293). The Medical Research Council (MRC) and the University of Bristol support the MRC Integrative Epidemiology Unit (MC_UU_00011/1). TC received funding from the European Union’s Horizon 2020 research and innovation programme under grant agreement No 733206, LIFE-CYCLE project. AH received funding from the Health Foundation’s Social and Economic Value of Health programme, grant number: 807293. LDH is supported by a Career Development Award from the UK Medical Research Council (MR/M020894/1). This work is part of a project entitled ‘social and economic consequences of health: causal inference methods and longitudinal, intergenerational data’, which is part of the Health Foundation’s Social and Economic Value of Health Programme (Grant ID: 807293). The Health Foundation is an independent charity committed to bringing about better health and health care for people in the UK. JALL was supported by the UK Economic and Social Research Council [grant number ES/P00881X/1]. Dr Wright is funded by a Cancer Research UK Population Research Postdoctoral Fellowship (C60153/A23895). GDS works in the Medical Research Council Integrative Epidemiology Unit at the University of Bristol [MC_UU_00011/1].

**ABBREVIATIONS**
ALSPAC: Avon Longitudinal Study of Parents and Children
UK: United Kingdom
IQ: Intelligence Quotient
RCT: Randomised Control Trial
GCSE: General Certificate of Secondary Education
CI: Confidence Interval
RMSEA: Root Mean Square Error of Approximation
SRMR: Standardised Root Mean Residual
CFI: Comparative Fit Index
TLI: Tucker Lewis Index

## Notes

### Competing Interest Statement

The authors have declared no competing interest.

### Author Declarations

Ethical approval for the study was obtained from the ALSPAC Ethics and Law Committee and the Local Research Ethics Committees.

